# Dynamics of seroconversion of anti-SARS-CoV-2 IgG antibodies in the Czech unvaccinated population: nationwide prospective seroconversion (PROSECO) study

**DOI:** 10.1101/2021.08.15.21262007

**Authors:** Pavel Piler, Vojtěch Thon, Lenka Andrýsková, Kamil Doležel, David Kostka, Tomáš Pavlík, Ladislav Dušek, Hynek Pikhart, Martin Bobák, Srdan Matic, Jana Klánová

## Abstract

**Background:** Although the Czech Republic weathered the first wave of the COVID-19 epidemic with relatively low incidence, the second wave of the global pandemic saw it rank among countries bearing the greatest COVID-19 burden, both in Europe and on a worldwide scale. The aim of the nationwide prospective seroconversion (PROSECO) study was to investigate the dynamics of seroconversion of anti-SARS-CoV-2 IgG antibodies in the Czech population.

**Methods:** All clients of the second largest health insurance company in the Czech Republic were sent a written invitation to participate in this longitudinal study. The study includes the first 30,054 persons who provided a blood sample between October 2020 and March 2021. Seroprevalence was compared between calendar periods of blood sample collection, RT-PCR test results, sociodemographic factors, and other characteristics.

**Findings:** The data show a dramatic increase in seropositivity over time, from 28% in October/November 2020 to 43% in December 2020/January 2021 to 51% in February/March 2021. These trends were consistent with government data on cumulative viral antigenic prevalence in the population captured by PCR testing – although the seroprevalence rates established in this study were considerably higher than those listed in government data. Data pooled across the entire study period exhibited minor differences in seropositivity between sexes, age groups and body mass index categories; results were similar between test providing laboratories. Seropositivity was substantially higher among symptomatic vs. asymptomatic persons (76% vs. 34%). At least one third of all seropositive participants were asymptomatic, and 28% participants who developed antibodies against SARS-CoV-2 never underwent PCR testing.

**Interpretation:** Antibody response provides a better marker of past SARS-CoV-2 infection than PCR testing data. Our data on seroconversion confirm the rapidly increasing prevalence in the Czech population during the dramatically rising pandemic wave prior to the beginning of massive vaccination. The planned second and third assessment of the study participants (April 2021 – September 2021, October 2021 – March 2022) will provide valuable evidence on the seroprevalence changes following vaccination and persistence of antibodies resulting from natural infection and vaccination.

**Research in context:** *Evidence before this study:* Similarly to most European countries, the first COVID-19 epidemic wave in the Czech Republic produced a relatively low incidence (86.9 confirmed cases per 100,000 persons over three months). At the peaks of the second wave, however, over 100 confirmed cases per 100,000 persons were diagnosed daily and the Czech Republic ranked among the countries with the greatest burden of COVID-19 in Europe and in the world. Only a few nationwide population-based studies have been published covering the second wave of the epidemic in Europe, and none of them from the Central and Eastern European region.

*Added value of this study:* The PROSECO study will provide key data from the heavily affected Central European region and contribute to the epidemiological and serological characteristics of the SARS-CoV-2 infection. All 30,054 study participants were recruited between October 2020 and March 2021, thus covering all three epidemic peaks (November 2020, January and March 2021) of the second COVID-19 epidemic wave. This allows us to follow the dynamics of seroconversion of anti-SARS-CoV-2 IgG antibodies in the immunologically naive and unvaccinated population during the COVID-19 pandemic. The study participants will be re-assessed in the second (April 2021 – September 2021) and third (October 2021 – March 2022) PROSECO phases to further study the post-infection/post-vaccination dynamics of seroconversion in/after a period of massive vaccination.

*Implications of all the available evidence:* Data from the first phase of the PROSECO study indicate that the percentage of the population that has been exposed to the SARS-CoV-2 may be substantially higher than estimates based on official data on cumulative viral positivity incidence as at least one third of seropositive participants were asymptomatic, and 28% of participants who developed antibodies against SARS-CoV-2 never underwent PCR testing. Regional seroprevalence data provide key information to inform, in combination with other surveillance data, public health policies and will be instrumental for the successful management of the subsequent phases of the global pandemic. The number of seropositive participants who never underwent RT-PCR testing demonstrates the importance of serological population-based studies describing the spread and exposure to the virus in the population over time.

## Introduction

The COVID-19 pandemic is caused by severe acute respiratory syndrome coronavirus 2 (SARS-CoV-2).^1^ This virus stimulates a rapid seroconversion to IgG antibodies in symptomatic^2,3^ as well as asymptomatic subjects.^4,5^ Therefore, the memory IgG antibodies against SARS-CoV-2 can serve as a specific long-term biomarker of previous SARS-CoV-2 infection^6^ as well as an appropriate marker for monitoring the persistence of antibodies against SARS-CoV-2 after vaccination.^7^

The SeroTracker dashboard, which systematically monitors and synthesises findings from hundreds of global SARS-CoV-2 serological studies^8^, shows a lack of nationwide population-based seroprevalence studies in Central and Eastern Europe, compared to several seroprevalence studies in other parts of Europe.^9-11^ While some existing studies have examined general population samples (ranging between 959^12^ and 105,651^13^ participants), most have been restricted to hotspots.^14-17^ These studies adopted different sampling and analytical strategies: some have used household and community sampling^9,17^ while others focused on biosamples from blood donors^18^ or residual sera in laboratories.^10,12^ While the majority of these studies employed serological tests, point-of-care antibody assays with lower sensitivity and specificity were also used.^13,19^ A review by Rostami et al. reported seroprevalence for Europe between 0.66% and 5.27% until August 2020.^20^ Thus far, only a small number of nationwide population-based studies have been published covering the period between November 2020 and March 2021, i.e. the second wave of the epidemic in Europe.^21-23^

In the Czech Republic, the first COVID-19 cases were confirmed on March 1, 2020. Due to successful government measures, the first epidemic wave in spring 2020 was relatively modest, with only 9,301 cases recorded by the end of May 2020 (86.9 confirmed cases per 100,000 persons over three months). However, the dismantling of restrictions in the summer resulted in a dramatic worsening of the epidemic situation in autumn 2020. At the peak of the second wave, over 10,000 new cases were diagnosed every day (over 100 confirmed cases per 100,000 persons daily) and the Czech Republic ranked among the countries with the greatest burden of COVID-19 in Europe and in the world. As of May 31, 2021, the Czech Republic had a cumulative total of 1,661,787 cases confirmed (15,528 per 100,000 persons) and the highest cumulative incidence of COVID-19 among countries with ≥1,000,000 inhabitants.^24^

To address the lack of regional data and to investigate the dynamics of seroprevalence of anti-SARS-CoV-2 IgG antibodies in the region, a nationwide prospective seroconversion study (PROSECO) was initiated in the Czech Republic. The aim was to study the dynamics of seroconversion in three six-month-long periods: 1) the second epidemic wave (unvaccinated population); 2) the mass vaccination period; and 3) the post-vaccination period. Here, we describe the study design and the results of the first phase of the study (October 2020 to March 2021).

## Methods

### Study design and study population

The Czech PROSECO study investigates seroconversion after SARS-CoV-2 infection (or vaccination), as well as the decline of memory IgG antibodies against SARS-CoV-2 over time. The study has been planned as a series of the three consecutive phases, each lasting six months, with participant enrolment taking place in the first phase. The first phase of the study, described here, was completed just before the launch of a massive nationwide vaccination programme.

All clients of the second largest health insurance company in the Czech Republic were sent a written invitation to participate in the study. On September 20 the first batch of invitation letters was sent to all clients (>18 years old) and the first 30,054 persons who provided blood samples between October 1, 2020 and March 31, 2021 were included in the study population. The presence of COVID-19 symptoms on the day of study enrolment was used as the sole exclusion criterion. The participants paid less than 20% (i.e. a negligible sum) of an antibody test cost.

The participants were invited to visit their local blood collection centres managed by one of the five different chains of clinical laboratories incorporated to QualityLab association covering the entire area of the Czech Republic. A 3 ml blood sample was collected in a separating gel vacuum tube. Serum samples were distributed at +4 °C and subsequently analysed in central laboratories within 12 hours. Leftover sera were stored in a biobank for future use.

Participants completed a questionnaire which included personal information (such as residence address, date of birth, sex), anthropometric data, self-reported results of previous RT-PCR tests (if performed), history of symptoms compatible with COVID-19, and records of COVID vaccination.

Informed consent forms were obtained from all study participants during each wave of the data collection. An ethics committee approval of all aspects of data collection, as well as of the secondary data analysis, was obtained from the ELSPAC ethics committee under reference number (C)ELSPAC/EK/5/2021.

### Detection of IgG antibodies against SARS-CoV-2

CE-marked serological tests were performed in accredited clinical laboratories. Antigen-specific humoral immune response was analyzed using commercial immunoassays LIAISON SARS-CoV-2 S1/S2 IgG (DiaSorin, Saluggia, Italy) and SARS-CoV-2 IgG II Quant (Abbott, Sligo, Ireland). Testing was conducted on the LIAISON XL (DiaSorin, Saluggia, Italy) and on the Alinity (Abbott, Lake Forest, IL, USA) respectively. Samples were tested individually and reported according to the manufactures’ criteria.

### Statistical analyses

The primary aim of this study was to determine the seroconversion rate of the adult Czech population. We estimated seroprevalence rates and 95% confidence intervals. We also standardized the seroprevalence rates to age and sex, using the Czech population as a standard.^25^ We used a multivariate Poisson regression model with a robust error variance to evaluate the differences in the seroprevalence rate between study periods and to compare SARS-CoV-2 antibody reactive individuals to non-reactive individuals. Differences in prevalence were expressed as prevalence rate ratios (PRRs). We used standard descriptive statistics to characterize the study data set.

Population data on COVID-19 were obtained from the Czech Central Information System of Infectious Diseases (ISID), which includes records of all consecutive patients with COVID-19 in the Czech Republic identified and confirmed by laboratory testing. ISID data are routinely collected in compliance with Act No. 258/2000 Coll. On the Protection of Public Health and are publicly available in aggregated and anonymized form of open or authenticated data sets. All analyses were conducted using Stata version 15.1 (StataCorp, College Station, Texas 77845 USA).

## Results

Between October 2020 and March 2021, 30,054 individuals consented to participate in the study and their results were included in the final analyses. The seroconversion overview is shown in Table 1. During the entire study period, 14,061 (46.8%) of the 30,054 individuals tested positive for anti-SARS-CoV-2 IgG antibodies. After adjusting for national population distribution with regard to sex and age, this corresponds to 46.4% (95% CI: 45.8–47.0). The seroconversion rate increased over time, along with the rise of the epidemic curve during the study period (Figure 1). In the first two months (October – November 2020), we estimated an overall seroconversion rate of 27.9% (95% CI: 26.1– 29.7). This estimate increased to 42.2% (95% CI: 40.8–43.5) in next two months (December 2020 to January 2021), and to 51.0% (95% CI: 50.3–51.8) in February and March 2021.

**Table 1:** Overview of seroconversion estimates between October 2020 and March 2021

| **Period** | **SARS-CoV-2 serology test result** |  | **Seroprevalence (95% CI) standandardized by age and sex for national population** | **p value** |
| --- | --- | --- | --- | --- |
|  | Total<br>n | Positive<br>n (%) |  |  |
| October 2020 – November 2020 | 3,626 | 1,025 (28.3%) | 27.9% (26.1–29.7) | p<0.001 |
| December 2020 – January 2021 | 6,880 | 2,984 (43.4%) | 42.2% (40.8–43.5) |  |
| February 2021 – March 2021 | 19,548 | 10,052 (51.4%) | 51.0% (50.3–51.8) |  |
| **Total** | **30,054** | **14,061 (46.8%)** | **46.4% (45.8–47.0)** |  |

**Figure 1:**
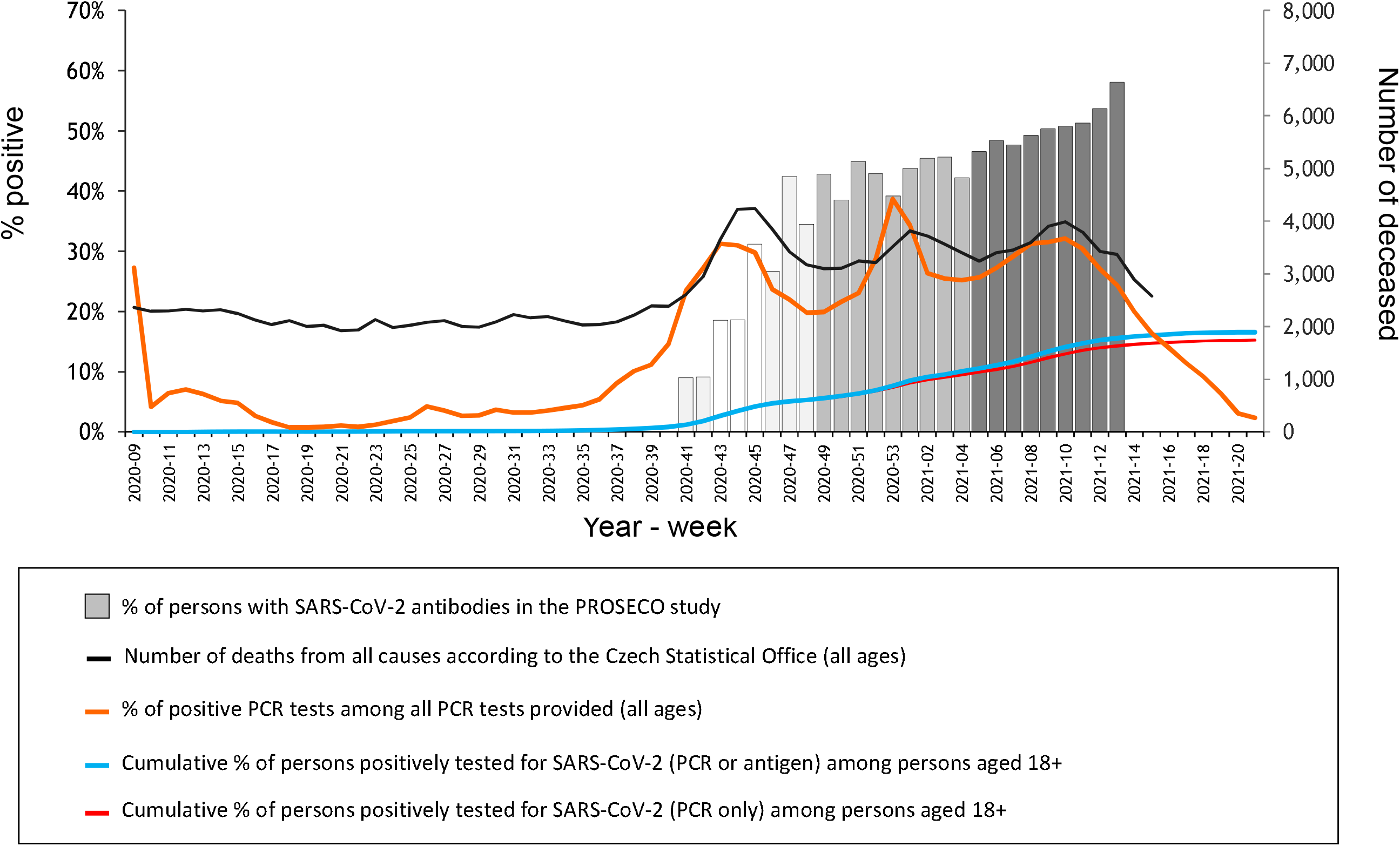
Dynamics of the COVID-19 pandemic in the Czech Republic and seroconversion in the first phase of the PROSECO study between October 2020 and March 2021

Table 2 shows differences in seroprevalence rates between various population groups. Seropositivity was slightly higher in the 18 to 29 and 50 to 59 age groups than among those aged 30 to 39, 40 to 49 and 60+. Rates were similar in men and women. Individuals who reported having previously one or more COVID-19 symptoms had higher seroprevalence when compared to asymptomatic participants (76.9% vs. 33.7%). A higher seropositivity prevalence was also observed for persons with BMI > 30 kg/m^2^. No major differences were established in seropositivity patterns with respect to age, sex, or the number of COVID-19 symptoms between the three two-month-long periods, or between the participating laboratories.

**Table 2:** Prevalence rate ratios (PRRs) and 95 % confidence intervals for seroprevalence of IgG antibodies to SARS-CoV-2 in PROSECO study participants estimated by multivariate Poisson regression

|  | October 2020 – November 2020<br>n = 3,626 |  |  |  | December 2020 – January 2021<br>n = 6,880 |  |  |  | February 2021 – March 2021<br>n = 19,548 |  |  |  | All study periods<br>n = 30,054 |  |  |  |
| --- | --- | --- | --- | --- | --- | --- | --- | --- | --- | --- | --- | --- | --- | --- | --- | --- |
|  | Particip.<sup>1</sup><br>n<br>(%) | Seropositive<sup>2</sup><br>(%) | PRR<br>(95% CI) | p-value | Particip.<sup>1</sup><br>n<br>(%) | Seropositive<sup>2</sup><br>(%) | PRR<br>(95% CI) | p-value | Particip.<sup>1</sup><br>n<br>(%) | Seropositive<sup>2</sup><br>(%) | PRR<br>(95% CI) | p-value | Particip.<sup>1</sup><br>n<br>(%) | Seropositive<sup>2</sup><br>(%) | PRR<br>(95% CI) | p-value |
| **Study periods** |  |  |  |  |  |  |  |  |  |  |  |  |  |  |  |  |
| 10/2020–11/2020 | - | - | - | - | - | - | - | - | - | - | - | - | 3,626<br>(12.1%) | 28.3% | 1 | - |
| 12/2020–01/2021 | - | - | - | - | - | - | - | - | - | - | - | - | 6,880<br>(22.9%) | 43.4% | 1.41<br>(1.34–1.49) | <0.001 |
| 02/2021–03/2021 | - | - | - | - | - | - | - | - | - | - | - | - | 19,548<br>(65.0%) | 51.4% | 1.67<br>(1.59–1.76) | <0.001 |
| **Sex** |  |  |  |  |  |  |  |  |  |  |  |  |  |  |  |  |
| Male | 1,424<br>(39.3%) | 27.7% | 1 | - | 2,756<br>(40.1%) | 41.8% | 1 | - | 7,612<br>(38.9%) | 51.1% | 1 | - | 11,792<br>(39.2%) | 46.1% | 1 | - |
| Female | 2,202<br>(60.7%) | 28.7% | 1.00<br>(0.90–1.11) | 0.996 | 4,124<br>(59.9%) | 44.4% | 1.04<br>(0.98–1.09) | 0.171 | 11,936<br>(61.1%) | 51.7% | 1.02<br>(0.99–1.04) | 0.223 | 18,262<br>(60.8%) | 47.2% | 1.02<br>(1.00–1.04) | 0.086 |
| **Age groups** |  |  |  |  |  |  |  |  |  |  |  |  |  |  |  |  |
| 18–29 | 318<br>(8.8%) | 31.1% | 1 | - | 582<br>(8.5%) | 43.1% | 1 | - | 1,660<br>(8.5%) | 52.5% | 1 | - | 2,560<br>(8.5%) | 47.7% | 1 | - |
| 30–39 | 662<br>(18.3%) | 24.8% | 0.80<br>(0.66–0.98) | 0.034 | 976<br>(14.2%) | 36.9% | 0.83<br>(0.74–0.94) | 0.002 | 2,621<br>(13.4%) | 47.9% | 0.90<br>(0.85–0.95) | <0.001 | 4,259<br>(14.2%) | 41.8% | 0.88<br>(0.83–0.92) | <0.001 |
| 40–49 | 1,354<br>(37.3%) | 28.3% | 0.91<br>(0.76–1.09) | 0.299 | 2,262<br>(32.9%) | 43.8% | 0.97<br>(0.88–1.07) | 0.548 | 5,515<br>(28.2%) | 51.4% | 0.95<br>(0.90–1.00) | 0.032 | 9,131<br>(30.4%) | 46.1% | 0.95<br>(0.91–0.99) | 0.020 |
| 50–59 | 842<br>(23.2%) | 31.0% | 0.98<br>(0.82–1.19) | 0.866 | 1,845<br>(26.8%) | 47.5% | 1.06<br>(0.96–1.17) | 0.219 | 5,020<br>(25.7%) | 53.7% | 0.99<br>(0.94–1.04) | 0.574 | 7,707<br>(25.6%) | 49.8% | 1.00<br>(0.96–1.05) | 0.884 |
| 60+ | 450<br>(12.4%) | 26.2% | 0.87<br>(0.70–1.08) | 0.218 | 1,215<br>(17.7%) | 41.6% | 0.97<br>(0.87–1.08) | 0.572 | 4,732<br>(24.2%) | 50.6% | 0.94<br>(0.90–1.00) | 0.032 | 6,397<br>(21.3%) | 47.2% | 0.95<br>(0.90–0.99) | 0.022 |
| **BMI categories** |  |  |  |  |  |  |  |  |  |  |  |  |  |  |  |  |
| Below 18.5 | 43<br>(1.2%) | 23.3% | 1 | - | 79<br>(1.1%) | 35.4% | 1 | - | 219<br>(1.1%) | 50.7% | 1 | - | 341<br>(1.1%) | 43.7% | 1 | - |
| 18.5–24.9 | 1,356<br>(37.4%) | 28.4% | 1.20<br>(0.73–1.96) | 0.466 | 2,568<br>(37.3%) | 41.5% | 1.11<br>(0.84–1.47) | 0.466 | 7,059<br>(36.1%) | 47.9% | 0.96<br>(0.85–1.08) | 0.500 | 10,983<br>(36.5%) | 44.0% | 1.00<br>(0.90–1.12) | 0.933 |
| 25.0–29.9 | 1,111<br>(30.6%) | 29.3% | 1.27<br>(0.77–2.10) | 0.340 | 2,414<br>(35.1%) | 45.0% | 1.19<br>(0.90–1.58) | 0.232 | 6,915<br>(35.4%) | 52.2% | 1.05<br>(0.90–1.18) | 0.442 | 10,440<br>(34.7%) | 48.1% | 1.09<br>(0.98–1.22) | 0.129 |
| 30.0 and above | 611<br>(16.9%) | 29.8% | 1.23<br>(0.74–2.04) | 0.423 | 1,316<br>(19.1%) | 47.3% | 1.23<br>(0.93–1.64) | 0.154 | 4,064<br>(20.8%) | 56.4% | 1.10<br>(0.98–1.24) | 0.118 | 5,991<br>(19.9%) | 51.7% | 1.13<br>(1.02–1.27) | 0.026 |
| Unknown | 505<br>(13.9%) | 24.2% | 1.25<br>(0.72–2.18) | 0.434 | 503<br>(7.3%) | 36.2% | 1.31<br>(0.95–1.79) | 0.097 | 1,291<br>(6.6%) | 50.7% | 1.12<br>(0.98–1.29) | 0.104 | 2,299<br>(7.7%) | 41.7% | 1.15<br>(1.02–1.27) | 0.025 |
|  | Particip.<sup>1</sup><br>n<br>(%) | Seropositive<sup>2</sup><br>(%) | PRR<br>(95% CI) | p-value | Particip.<sup>1</sup><br>n<br>(%) | Seropositive<sup>2</sup><br>(%) | PRR<br>(95% CI) | p-value | Particip.<sup>1</sup><br>n<br>(%) | Seropositive<sup>2</sup><br>(%) | PRR<br>(95% CI) | p-value | Particip.<sup>1</sup><br>n<br>(%) | Seropositive<sup>2</sup><br>(%) | PRR<br>(95% CI) | p-value |
| **COVID symptoms** |  |  |  |  |  |  |  |  |  |  |  |  |  |  |  |  |
| Asymptomatic | 2,266<br>(62.5%) | 18.1% | 1 | - | 3,941<br>(57.3%) | 29.4% | 1 | - | 11,119<br>(56.9%) | 38.5% | 1 | - | 17,326<br>(57.7%) | 33.7% | 1 | - |
| Symptomatic | 767<br>(21.2%) | 62.3% | 3.47<br>(3.13–3.85) | <0.001 | 2,080<br>(30.2%) | 75.0% | 2.61<br>(2.47–2.76) | <0.001 | 5,873<br>(30.0%) | 79.5% | 2.09<br>(2.03–2.15) | <0.001 | 8,720<br>(29.0%) | 76.9% | 2.27<br>(2.22–2.33) | <0.001 |
| Unknown | 593<br>(16.4%) | 23.1% | 1.21<br>(0.91–1.60) | 0.191 | 859<br>(12.5%) | 30.8% | 1.11<br>(0.97–1.28) | 0.139 | 2,556<br>(13.1%) | 43.3% | 1.17<br>(1.10–1.25) | <0.001 | 4,008<br>(13.3%) | 37.6% | 1.17<br>(1.11–1.24) | <0.001 |
| **Test provider** |  |  |  |  |  |  |  |  |  |  |  |  |  |  |  |  |
| Test provider 1 | 143<br>(3.9%) | 22.4% | 1 | - | 241<br>(3.5%) | 29.9% | 1 | - | 520<br>(2.7%) | 51.7% | 1 | - | 904<br>(3.0%) | 41.3% | 1 | - |
| Test provider 2 | 1,517<br>(41.8%) | 28.3% | 0.94<br>(0.64–1.39) | 0.769 | 2,745<br>(39.9%) | 44.8% | 1.16<br>(0.90–1.51) | 0.257 | 8,100<br>(41.4%) | 54.3% | 0.97<br>(0.86–1.09) | 0.546 | 12,362<br>(41.1%) | 49.0% | 1.02<br>(0.91–1.13) | 0.765 |
| Test provider 3 | 623<br>(17.2%) | 29.9% | 1.10<br>(0.77–1.58) | 0.602 | 1,212<br>(17.6%) | 43.0% | 1.13<br>(0.88–1.45) | 0.349 | 3,495<br>(17.9%) | 48.5% | 0.85<br>(0.76–0.95) | 0.004 | 5,330<br>(17.7%) | 45.0% | 0.92<br>(0.83–1.02) | 0.111 |
| Test provider 4 | 1,182<br>(32.6%) | 29.1% | 1.08<br>(0.73–1.59) | 0.701 | 2,426<br>(35.3%) | 42.9% | 1.29<br>(1.00–1.68) | 0.054 | 6,703<br>(34.3%) | 49.3% | 0.96<br>(0.85–1.08) | 0.463 | 10,311<br>(34.3%) | 45.5% | 1.04<br>(0.94–1.16) | 0.453 |
| Test provider 5 | 161<br>(4.4%) | 20.5% | 0.69<br>(0.43–1.10) | 0.121 | 256<br>(3.7%) | 46.5% | 1.16<br>(0.88–1.53) | 0.295 | 730<br>(3.7%) | 52.5% | 0.91<br>(0.80–1.03) | 0.130 | 1,147<br>(3.8%) | 46.6% | 0.95<br>(0.84–1.06) | 0.346 |

Particip.<sup>1</sup> = Number of participants; Seropositive<sup>2</sup> = Percentage of seropositive participants; PRR = Prevalence rate ratio; COVID symptoms = Symptoms compatible with COVID-19

Table 3 shows that RT-PCR tests were previously performed on 15,799 individuals (52.6% of the study sample) and of these, 62.0% were found SARS-CoV-2 seropositive. Seropositivity was also detected in 28.4% of individuals who were never tested using RT-PCR and in 29.3% of those who reported negative RT-PCR test results. On the other hand, 15.8% of the individuals reporting positive RT-PCR test results were found seronegative.

**Table 3:** SARS-CoV-2 seroprevalence based on RT-PCR status

|  | **Number of seropositive**<br>**participants**<br>**n (%)** | **Number of seronegative**<br>**participants**<br>**n (%)** | **All participants**<br>**n (%)** |
| --- | --- | --- | --- |
| **RT-PCR status (n=30,054)** |  |  |  |
| Never performed | 3,554 (28.4%) | 8,950 (71.6%) | 12,504 (100%) |
| Performed | 9,799 (62.0%) | 6,000 (38.0%) | 15,799 (100%) |
| Unknown | 708 (40.4%) | 1,043 (59.6%) | 1,751 (100%) |
| **RT-PCR results (n=15,799)** |  |  |  |
| Negative | 1,736 (29.3%) | 4,199 (70.7%) | 5,935 (100%) |
| Positive | 7,846 (84.2%) | 1,475 (15.8%) | 9,321 (100%) |
| Unknown | 217 (40.0%) | 326 (60.0%) | 543 (100%) |

Figure 1 shows the weekly seroprevalence in this study along with national cumulative data on SARS-CoV-2 positivity (blue line). While the absolute levels are very different, the temporal trends established using the two data sources are virtually identical, with a very rapid increase (over five-fold) between October 2020 and March 2021. Both positive PCR tests (orange line) as well as cumulative PCR values (red/blue line) demonstrate the rapid spread of the virus in the Czech population, which resulted in a dramatic increase in all-cause mortality throughout this period.

## Discussion

This study demonstrated a rapid increase in anti-SARS-CoV-2 IgG antibodies in the unvaccinated Czech adult population during the second COVID-19 pandemic wave between October 2020 and March 2021. We found a massive dynamic increase in seroconversion rate of IgG antibodies against SARS-CoV-2 from 9% to 51.0% during the course of six months (Figure 1).

Seroprevalence was low at the beginning of the study (October 2020), which is in agreement with similar studies performed in other countries until September 2020. Data available for a similar period from the Netherlands, France, Spain, UK, and Italy, for instance, suggested seroprevalence oscillating between 3.4% and 11.6%.^9,10,12,13,18^ By the end of March 2021, the seroconversion rate reached 50% in our study subjects. The dynamics of seroprevalence in our study corresponded with nationwide Czech government data.

Only small differences in the seroprevalence of anti-SARS-CoV-2 IgG antibodies were observed among various population groups. Similarly to previous studies^9,14,26^, no difference was found between men and women (Table 2). In agreement with other publications, our results indicated some differences in prevalence between age groups.^26,27^ The lowest rate of seropositivity was observed in the 30–39 age group, followed by the 40–49 and 60+ age groups. The highest risk was estimated for the youngest age group 18–29 and for people in their fifties (50–59). We can only hypothesize that many people in their thirties were parents of small children who could not attend school for a majority of the studied period. This may have thus led to their more efficient isolation in case they were able to take advantage of paid leave or home office schemes. Simultaneously, a majority of this 30–39 age group was probably still in good health and not suffering from chronic conditions. As the youngest age group has been generally regarded as the one with the lowest respect for the disease and related restrictions, it is not surprising that their risk is rather high. They can be expected to have many more contacts than other sub-groups but also the smallest percentage of individuals with chronic conditions. The highest seropositivity rate was found in the 50–59 age group. Most people in this group are still actively working and many of them were present in their workplaces for majority of the study period (with the exception of several weeks when the most stringent restrictions were in place and a majority of businesses and offices were closed). They may also be less apt at using IT technologies than the younger age groups and thus benefit less from home office options. At the same time, a higher percentage of these individuals is expected to suffer from various chronic conditions. As demonstrated in previously published studies such as Ward et al. in the UK^13^, we also established a higher risk for the obese sub-population.

Most seropositive participants reported more than one symptom related to the SARS-CoV-2 infection. On the other hand, around 30% of seropositive subjects were asymptomatic, which is consistent with other studies.^9,13^ The ratio of asymptomatic and symptomatic participants differs among study phases: twice as many seropositive participants were asymptomatic in the third vs. in the first phase. This could be explained by exposure to low doses of the virus repeatedly over time, which could lead to seroconversion without the manifestation of symptoms (Table 2).

It should be also noted that 28% of participants who never underwent RT-PCR testing for SARS-CoV-2 were seropositive. Furthermore, 29% of seropositive participants who were tested by RT-PCR had previously had negative RT-PCR test results. Neither of these groups were visible in the official government statistics. On the other hand, 16% of seronegative participants previously reported a positive PCR test (Table 3). It may be expected that the mucosa-associated lymphoid tissue of these individuals dealt with the infection without triggering a systemic immune response.^28^

To the best of our knowledge, the Czech PROSECO study is the largest study of the seroconversion of anti-SARS-CoV-2 IgG antibodies in Central and Eastern Europe. According to the WHO seroepidemiological investigation protocol^29^, the major strengths of our study are its size and coverage, its launch prior to the beginning of the vaccination period and the on-going longitudinal follow-up. In contrast to other larger studies, we assessed antibody levels in the harmonized QualityLab network of accredited clinical laboratories and applied highly sensitive and specific chemiluminescence serological immunoassays targeting the receptor-binding domain (RBD) of the spike protein of SARS-CoV-2 virus. Since previous studies have shown that the SARS-CoV-2 spike protein RBD is the target of neutralizing antibodies, this molecule has become a particularly interesting target for serological testing and vaccine development.^30-32^ Our study has the advantage of including data from serological testing as well as self-reported RT-PCR test results. This allows us to more precisely explore the spread of infection in the nationwide study population.

The study also has important limitations. The main weakness is the self-selection of study participants. Potential participants were contacted through their health insurance company (newsletter, letter) and using online advertising. Similarly to other national seroprevalence studies, child populations was excluded for obvious reasons (informed consent, biological tissue requirement). The first 30,054 volunteers reporting to the QualityLab network were then enrolled in the study, after which the recruitment procedure was discontinued. As a result, the true response rate cannot be determined and, despite its large size, this study sample may not reliably represent the entire population, since the participants were self-selected volunteers rather than a random sample selected from a specific sampling frame. It is likely that persons at higher risk of past or current infection are overrepresented in this study, which would lead to an overestimation of the absolute seropositivity prevalence rate. Crucially, however, the temporal trend in seropositivity in this study sample closely reflects the cumulative PCR positivity trend in national data, thus suggesting the good internal validity of our results.

Compared to the national statistics, female and middle-aged participants (40 to 59 years) were more represented in our study at the expense of males, respondents from 18 to 39 years, and those over 60 years of age. The discrepancy in age profile can be partially attributed to the fact that the study participants were clients of one particular health insurance company, which has a larger representation of middle-aged clients compared to the general population. We also excluded the institutionalised population, which resulted in an underestimation of the oldest age category living in retirement/nursing homes, and individuals who were in prisons or monasteries at the time of the study.

In March 2021, the national vaccination program started, aiming at vaccinating the entire population against SARS-CoV-2 by the end of September 2021. The second phase of the PROSECO study was therefore launched to follow up on the study participants throughout the most intense vaccination period (i.e. from April to September 2021). The third phase, planned from October 2021 to March 2022, will investigate the dynamics and persistence of IgG SARS-CoV-2 antibodies. This longitudinal follow-up, which thus includes pre-vaccination, vaccination and postvaccination periods, will further increase importance and impact of this surveillance.^33^

In conclusion, the results of the first phase of the PROSECO study indicate that a significant part of the Czech adult population is no longer immunologically naïve to the SARS-CoV-2 infection. The massive increase of the seroconversion rate demonstrates the extensive exposure of the Czech population to the SARS-CoV-2 virus between October 2020 and March 2021. The number of seropositive participants who never underwent RT-PCR testing demonstrates the importance of serological population-based studies describing the spread and exposure to the virus in the population over time.

## Data Availability

no available data

## Contributors

VT, PP, LA and JK were responsible for the design of the study, LA for ethics, and DK, LA and VT for communication to potential participants. KD, PP and LA were responsible for the study operation, coordination of data acquisition and logistics, and KD for coordination and quality management of participating laboratories. TP, PP and LD were in charge of statistical analyses and table and figure design. The first draft was written by PP and VT, MB, HP and SM provided expertise in epidemiology. All authors contributed to data interpretation, critically reviewed the first draft, approved the final version and agreed to be accountable for the work.

## Data sharing

Our data are accessible to researchers upon reasonable request for data sharing to the RECETOX Research Infrastructure.

## Acknowledgement

The PROSECO study was sponsored by the Prevention Programme of the Health Insurance Company of the Ministry of the Interior of the Czech Republic. The RECETOX Research Infrastructure was supported by the Ministry of Education, Youth and Sports of the Czech Republic (LM2018121), and VT and PP from the CETOCOEN PLUS project of ESIF (CZ.02.1.01/0.0/0.0/15_003/0000469). The authors acknowledge funding from the CETOCOEN EXCELLENCE Teaming Phase 2 project supported by Horizon 2020 (857560) and the Ministry of Education, Youth and Sport of the Czech Republic/ESIF (CZ.02.1.01/0.0/0.0/17_043/0009632. We thank all collaborating nurses, laboratories from the QualityLab association, and administrative personnel and especially the 30,054 participants who invested their time and provided samples and information for this study.

